# Evaluating the accuracy of different respiratory specimens in the laboratory diagnosis and monitoring the viral shedding of 2019-nCoV infections

**DOI:** 10.1101/2020.02.11.20021493

**Authors:** Yang Yang, Minghui Yang, Chenguang Shen, Fuxiang Wang, Jing Yuan, Jinxiu Li, Mingxia Zhang, Zhaoqin Wang, Li Xing, Jinli Wei, Ling Peng, Gary Wong, Haixia Zheng, Weibo Wu, Mingfeng Liao, Kai Feng, Jianming Li, Qianting Yang, Juanjuan Zhao, Zheng Zhang, Lei Liu, Yingxia Liu

## Abstract

**Background:** The outbreak of novel coronavirus pneumonia (NCP) caused by 2019-nCoV spread rapidly, and elucidating the diagnostic accuracy of different respiratory specimens is crucial for the control and treatment of this disease.

**Methods:** Respiratory samples including nasal swabs, throat swabs, sputum and bronchoalveolar lavage fluid (BALF) were collected from Guangdong CDC confirmed NCP patients, and viral RNAs were detected using a CFDA approved detection kit. Results were analyzed in combination with sample collection date and clinical information.

**Findings:** Except for BALF, the sputum possessed the highest positive rate (74.4%∼88.9%), followed by nasal swabs (53.6%∼73.3%) for both severe and mild cases during the first 14 days after illness onset (d.a.o). For samples collected ≥ 15 d.a.o, sputum and nasal swabs still possessed a high positive rate ranging from 42.9%∼61.1%. The positive rate of throat swabs collected ≥ 8 d.a.o was low, especially in samples from mild cases. Viral RNAs could be detected in all the lower respiratory tract of severe cases, but not the mild cases. CT scan of cases 02, 07 and 13 showed typical viral pneumonia with ground-glass opacity, while no viral RNAs were detected in first three or all the upper respiratory samples.

**Interpretation:** Sputum is most accurate for laboratory diagnosis of NCP, followed by nasal swabs. Detection of viral RNAs in BLAF is necessary for diagnosis and monitoring of viruses in severe cases. CT scan could serve as an important make up for the diagnosis of NCP.

**Funding:** National Science and Technology Major Project, Sanming Project of Medicine and China Postdoctoral Science Foundation.

## INTRODUCTION

The outbreak of NCP caused by the novel coronavirus, designated as 2019-nCoV, started in Wuhan, China, at the end of 2019 ^1^. As of Feb. 5, 2020, at least 24324 cases and 490 deaths have been identified across China and other countries ^2-5^. On Jan. 30, 2020, WHO has declared that the outbreak of 2019-nCoV constitutes a Public Health Emergency of International Concern (PHEIC) and issued this advice as temporary recommendations under the International Health Regulations (IHR).

Clinical features varied in different cases, and some patients showed asymptomatic infection ^2,6-8^. Recent studies have confirmed the human to human transmission of 2019-nCoV ^2,3,6,8^. More importantly, asymptomatic cases could transmit virus to other contacts, which makes it more difficult to control the spread of the virus ^2^. Rapid and accurate detection of 2019-nCoV is in urgent need due to the rapid spread and increasing number of NCP patients ^6^. Studies have shown that pneumonia is the common complication of 2019-nCoV infection ^6,7^, which suggests that it mainly infects the lower respiratory tract. Until recently, the diagnosis of the cases in the published studies was mostly from the lower respiratory tract specimens ^7^. However, collection of the lower respiratory samples (usually BALF) requires both a suction device and a skilled operator, also painful for the patients. So, BALF samples are not feasible for the routine laboratory diagnosis and monitoring of the 2019-nCoV. Instead, collection of a nasal swab, throat swab and sputum is rapid, simple and safe.

Accordingly, elucidating the diagnosis accuracy of different sample types is crucial for the laboratory diagnosis and monitoring the viral shedding of 2019-nCoV. Moreover, no data on the difference of viral shedding between the upper and lower respiratory tract specimens is currently available. In this study, we aim to investigate the diagnostic accuracy of the upper respiratory samples, and compare the viral distribution and shedding between the mild and severe cases. We believe our results would help the laboratory staff and physicians in the diagnosis and treatments of patients with 2019-nCoV.

## METHODS

### Patients and samples

213 Guangdong CDC (Center for Disease Control and Prevention) confirmed 2019-nCoV infected patients who were hospitalized in Shenzhen Third People’s hospital between Jan 11 and Feb. 03, 2020 were included. A total of 866 samples from respiratory tracts of the patients including nasal swabs, throat swabs, sputum and BALF were collected upon admission and various time-points thereafter. Sample collection dates were divided into 0∼7, 8∼14 and ≥ 15 d.a.o groups, and patients were divided into severe and mild cases according to the guidelines of 2019-nCoV infection from the National Health Commission of the People’s Republic of China. The study was approved by the Ethics Committees from Shenzhen Third People’s Hospital (SZTHEC2016001).

### Quantitative reverse transcription polymerase chain reaction

Viral RNAs were extracted from the samples using the QIAamp RNA Viral Kit (Qiagen, Heiden, Germany), and quantitative reverse transcription polymerase chain reaction (qRT-PCR) was performed using a China Food and Drug Administration (CFDA) approved commercial kit specific for 2019-nCoV detection (GeneoDX Co., Ltd., Shanghai, China). The specimens were considered positive if the Ct value was ≤ 37.0, and negative if the results were undetermined. Specimens with a Ct higher than 37 were repeated. The specimen was considered positive if the repeat results were the same as the initial result and between 37 and 40. If the repeat Ct was undetectable, the specimen was considered negative.

### Quantification of hypoxia and lung injury

Quantification of hypoxia and lung injury was carried out as previously reported _9,10_. In brief, the partial pressure of oxygen (PaO_2_) in arterial blood taken from the patients at various time-points after hospitalization was measured by the ABL90 blood gas analyzer (Radiometer). The fraction of inspired oxygen (FiO_2_) is calculated by the following formula: FiO_2_ = (21 + oxygen flow (in units of l/min) × 4) / 100. The PaO_2_/FiO_2_ ratio (in units of mmHg) is calculated by dividing the PaO_2_ value with the FiO_2_ value.

### Statistics analyses

The unpaired, two-tailed *t*-test was used to determine whether differences in the Ct values were statistically significant. The Fisher exact test analysis was used to analyze positive rate. A *p*-value lower than 0.05 was considered statistically significant. Statistical analyses were performed using GraphPad Prism.

## RESULTS

### Patients and sample profile

Altogether, 866 respiratory specimens from 213 patients were collected, including 205 throat swabs, 490 nasal swabs, 142 sputum and 29 BALF. Of these patients, 37 were in severe or critical conditions, and the rest were mild cases (Table 1). The median age of severe cases is 65, and ranged from 34 to 81. Most of the patients belonged to the 45-64 (43.2%) and ≥ 65 (51.4) years of age groups (Table 1). For the mild cases, the median age is 47 with a range of 2 to 86. Unlike the severe cases, most of the mild cases belonged to the 15-44 (42.05%) and 45-64 (42.05%) years of age groups. The male ratio (62.2%) seems like higher than female in severe cases, but there is no statistical difference. Moreover, the sex ratio is similar in both groups of severe and mild cases. The median d.a.o of collection of the first specimen were 7 and 4 for the severe and mild cases, respectively (Table 1). The median number of specimens collected from each patient was 3 (range 1-23).

**Table 1.** Baseline characteristics and specimens of NCP cases.

| Characteristic | NCP cases |  |  |
| --- | --- | --- | --- |
|  | Total (N=213) | Severe (N=37) | Mild (N=176) |
| **Median age (range)** | 52 (2-86) | 65 (34-81) | 47 (2-86) |
| **Age subgroup (N, %)** | 213 | 37 | 176 |
| <15 yr | 9 (4.2) | 0 (0) | 9 (5.1) |
| 15-44 yr | 76 (35.7) | 2 (5.4) | 74 (42.05) |
| 45-64 yr | 90 (42.3) | 16 (43.2) | 74 (42.05) |
| ≥65 yr | 38 (17.8) | 19 (51.4) | 19 (10.8) |
| **Male (n, %)** | 108 (50.7) | 23 (62.2) | 85 (48.3) |
| **Sample types (N)** | 866 | 260 | 606 |
| Throat swabs | 205 (23.7) | 93 | 112 |
| Nasal swabs | 490 (56.6) | 96 | 394 |
| Sputum | 142 (16.4) | 45 | 97 |
| BALF | 29 (3.3) | 26 | 3 |
| **Median d.a.o of first specimen collection (range)** | 5 (1-17) | 7 (2-16) | 4 (1-17) |
| **Median number of specimens for each patient (range)** | 3 (1-23) | 5 (1-23) | 3 (1-12) |
| 0~7 d.a.o | 2 (1-7) | 2 (1-6) | 2 (1-7) |
| 8~14 d.a.o | 3 (1-10) | 4 (1-10) | 2 (1-9) |
| ≥15 d.a.o | 3 (1-16) | 5.5 (1-16) | 2 (1-6) |

d.a.o: Days after illness onset.
NCP: Novel coronavirus pneumonia.
yr: Years of age.

### Detection of 2019-nCoV in different respiratory sites NCP cases

The collected different types of specimens from 2019-nCoV confirmed cases were divided into three groups based on the collection time, including the 0∼7 d.a.o, 8∼14 d.a.o and ≥ 15 d.a.o groups. The qPCR assay was performed for each specimen and the results were shown in table 2. For the 0∼7 d.a.o group, the sputum sample showed the highest positive rate in both severe (88.9%) and mild (82.2%) cases, follow by nasal swabs (73.3%, 72.1%) and then the throat swabs (60.0%, 61.3%). In the mild cases, the positive rates from nasal swabs and sputum were similar. BLAF collected during 8∼14 d.a.o in the severe cases showed 100% positive, while negative in the BALF of three mild cases. The sputum collected during 8∼14 d.a.o also show the highest positive rate among the upper respiratory samples in both severe and mild cases, much higher than the nasal and throat swabs. Of note, the positive rate of throat swabs is only 50% in severe and 29.6% in mild cases. BALF, sputum and nasal swabs collected from severe cases ≥ 15 d.a.o showed similar positive rate. In mild cases, the positive rates of sputum and nasal swabs were similar, much higher than the throat swabs. In addition, the Ct values were significantly lower in sputum from severe cases.

**Table 2.**
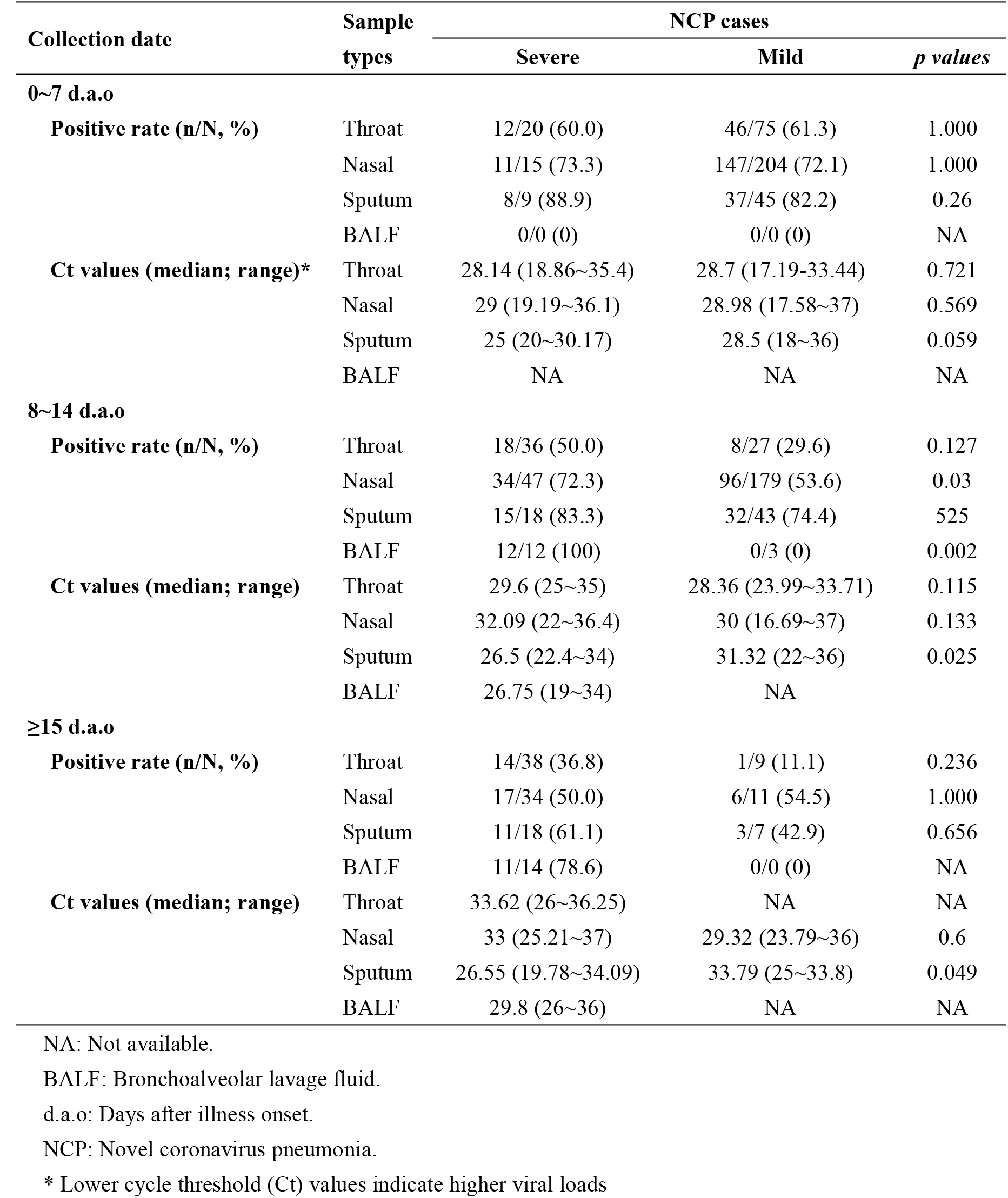
Detection of 2019-nCoV in respiratory sites of NCP cases.

### Profiles of viral shedding in severe and mild NCP cases

Serial samples from both upper (including throat swabs, nasal swabs and sputum, marked in red) and lower (BALF, marked in blue) respiratory tract from 13 NCP cases were collected and analyzed. The patients were grouped into severe (N=10, marked in red) and mild (N=3, marked in blue) cases and the detection results were shown in Figure 2. Viral RNAs could be detected in the upper respiratory tract samples collected during 3 and 21 d.a.o, and detected in BLAF at 23 d.a.o with high viral load. In severe cases, viral RNAs were detected in all the BALF samples as early as 6 d.a.o, and upper respiratory samples from 10 (10/11) cases. In case 2, although the Ct value was low in BALF, viral RNAs were not detected in all the upper respiratory samples. Meanwhile, for some severe cases (cases 06 and 07), viral RNAs were not detected in all the upper respiratory samples. As to the 3 mild cases, the viral RNAs was only detected in the upper respiratory samples, not in the BLAF. Moreover, the duration of viral shedding is longer in most of the severe cases.

**Figure 1.**
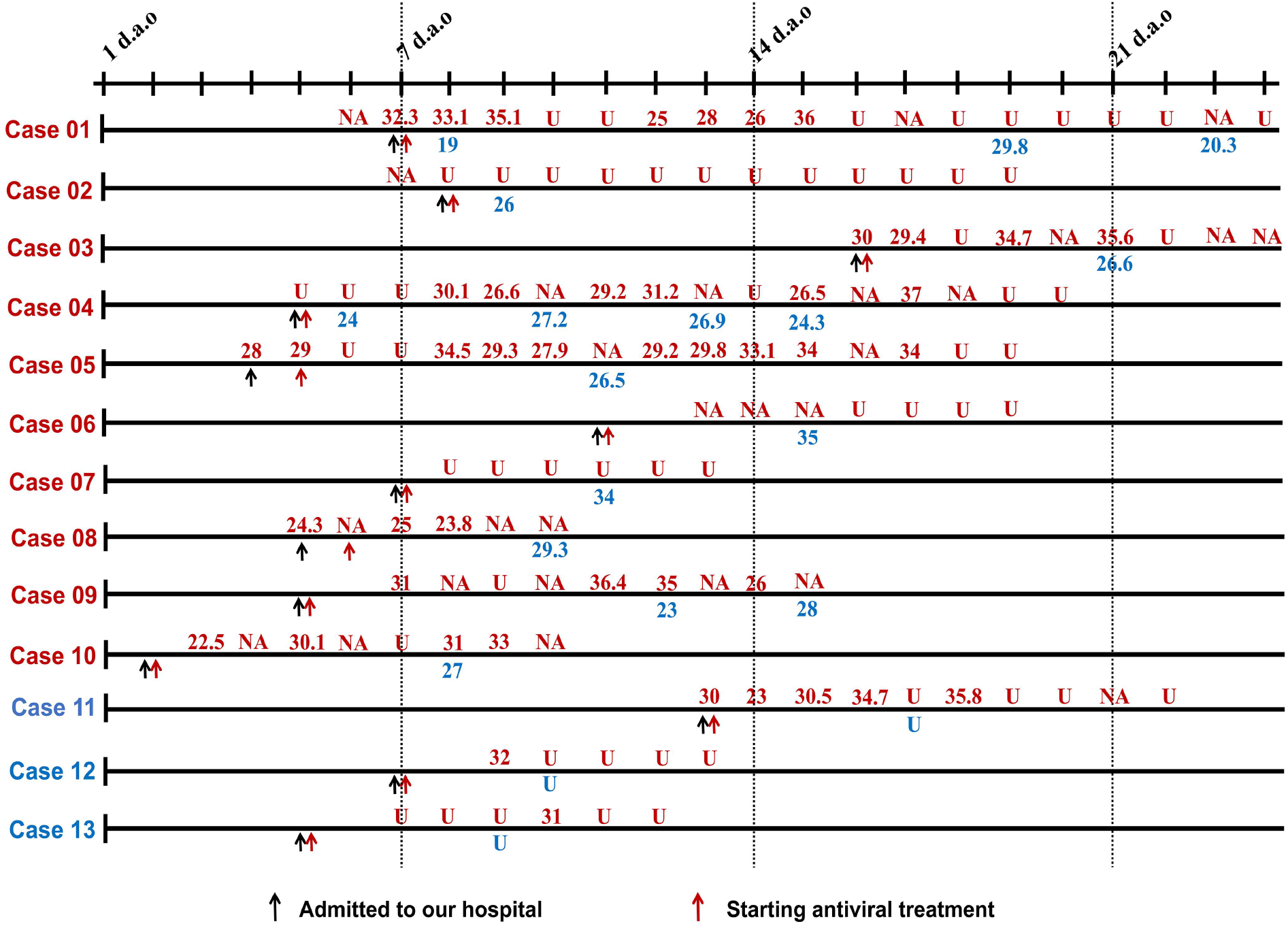
Serial detection of viral RNAs in the upper and lower respiratory tract of 13 NCP cases. Number of cases with severe condition were marked in red, and mild condition in blue. The detection results of samples from upper respiratory tract were in red, and lower respiratory tract in blue. Lower cycle threshold (Ct) values indicate higher viral loads.

**Figure 2.**
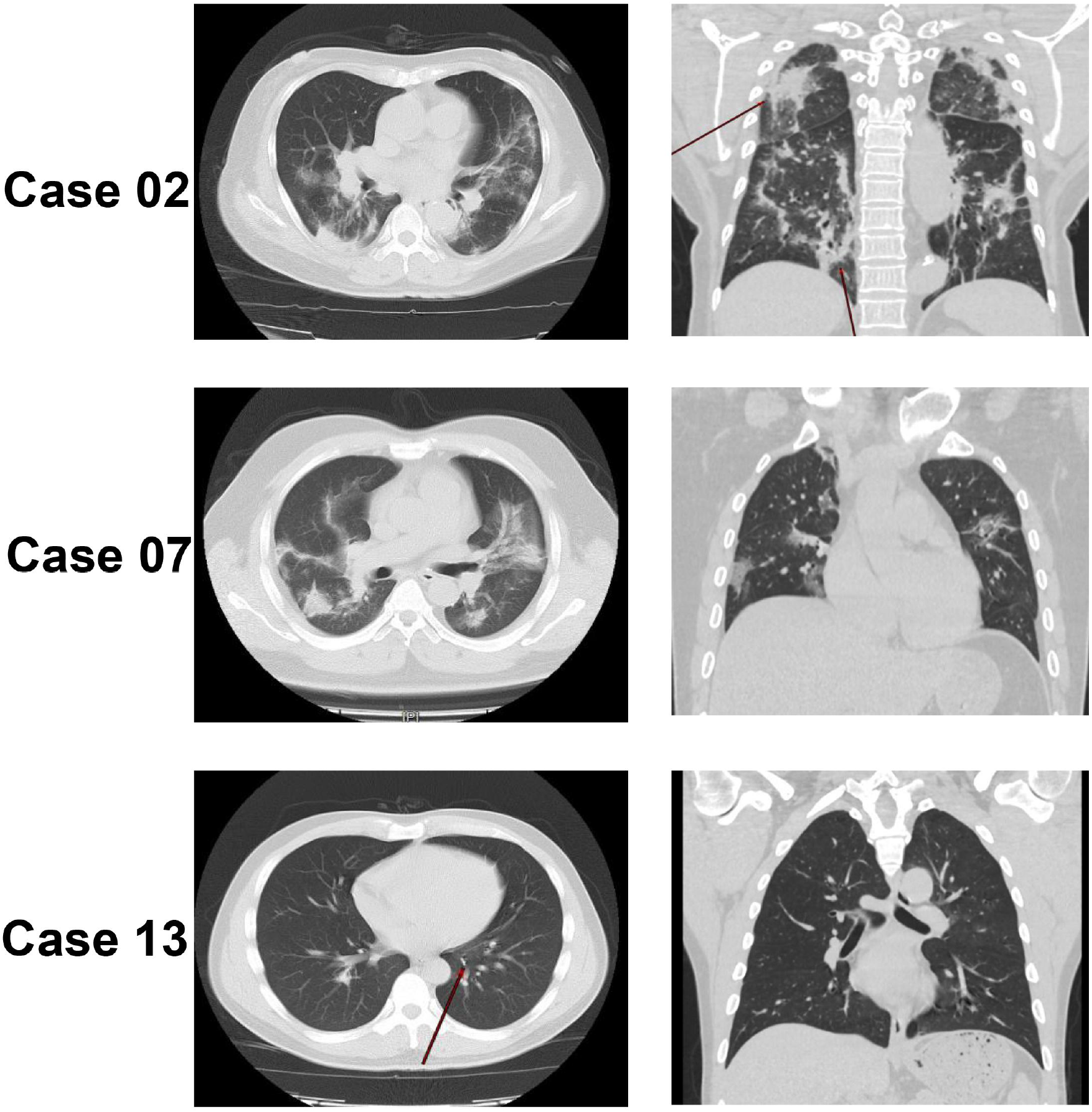
Computed tomography (CT) scan of the cases 02, 07 and 13.

### Computed tomography (CT) scan may serve as an important make up for the diagnosis of NCP

The epidemiological and clinical features of cases 02, 07 and 13 from whom viral RNA were not detected in the first three or all the upper respiratory samples were analyzed in detail (Table 3 and Fig 2). Case 02 was a female aged 65 with hypertension, and case 07 was a male aged 34 without any underling diseases. Both them had a travelling history to Wuhan. The CT scan of the two cases showed multiple ground-glass opacities in bilateral lungs. The PiO_2_/FiO_2_ and Murray score were 188 and 2 for case 02, 306 and 1.5 for case 07, which indicated a lung injury. For the two cases, no viral RNAs were detected in the upper respiratory tract but positive in the BALF. Cases 13 was a male 36 without any underling diseases. Due to the exposure history ^8^, CT scan and viral screen were done. The CT scan of the this case also showed typical ground-glass opacity in the lung, suggesting a viral pneumonia (Fig 2). However, no viral RNAs were detected until the fourth upper respiratory samples.

**Table 3.** Epidemiological and clinical characteristics of cases 2, 7 and 13.

| Case No. | Sex | Age | Initial symptoms | Underling diseases | Possible exposure | Indexes of lung injury |  |
| --- | --- | --- | --- | --- | --- | --- | --- |
|  |  |  |  |  |  | PiO<sub>2</sub>/FiO<sub>2</sub> | Murray score |
| Case 02 | Female | 65 | Fever, cough, myalgia, chill and diarrhea | Hypertension | Travelled to Wuhan | 188 | 1.75 |
| Case 07 | Male | 34 | Fever, myalgia and diarrhea | No | Lived in Wuhan | 306 | 1.5 |
| Case 13 | Male | 36 | Cough and diarrhea | No | Travelled to Wuhan with case 02 | 438 | 0.25 |

## DISCUSSION

According to our results, apart from the BALF collected during 8∼14 d.a.o which possessed the 100% (12/12) positive rate, sputum samples showed the highest positive rate in all stages post 2019-nCoV infection, and followed by nasal swabs. The positive rate of throat swab varied in the severe and mild cases. For the severe cases, the positive rates were similar in samples collected 0∼7 and 8∼14 d.a.o, while low in samples collected ≥ 15 d.a.o. For the mild cases, it showed the highest positive rate in samples collected 0∼7 d.a.o, however, very low positive rate in samples collected 8∼14 and ≥15 d.a.o. The results indicate that sputum may serve as the most sensitive samples for the virus detection, and followed by nasal swabs. However, a recent study found that only a small portion (28%) of NCP cases showed sputum production ^7^. As a result, nasal swabs may be the most widely applicable samples for virus detection. On the contrary, throat swabs were not recommended for the viruses detection, especially the samples collected 8∼14 and ≥ 15 d.a.o from mild cases, which may result in a large proportion of false negative results.

Laboratory detection of viral RNA in the respiratory samples of suspected individuals is now considered one of the criteria for the diagnosis of NCP, and the samples from upper respiratory tract were regularly used (http://www.nhc.gov.cn/yzygj/s7653p/202002/3b09b894ac9b4204a79db5b8912d4440.shtml). However, as shown in our study (Fig 1), viral RNAs could not be detected in the upper respiratory samples from some severe cases (cases 02, 06 and 07), while positive in the BALF. Moreover, in some patients like cases 04 and 13, the viruses were not detected in the first three samples, mostly in the 0∼7 d.a.o. The results suggest that the suspected patients especially those with exposure history and clinical symptoms might not be excluded from NCP despite that viral RNA was not detected in the upper respiratory samples. Since human to human transmission of 2019-nCoV have been proved in recent studies ^2,3,6,8^,we must pay more attention to these people, in case of further spread of the virus. Under such circumstances, CT scan might provide important make up for the diagnosis of NCP patients. For example, although no viral RNAs were detected in the first three or all the upper respiratory samples from cases 02, 07 and 13, the CT scans showed typical viral pneumonia linked to NCP ^7,11,12^, and finally 2019-nCoV were identified. Another notification is that, during the antiviral treatment, even though we did not detect the viral RNA in the upper respiratory tract, while it was still positive in the BALF samples of some patients (cases 01, 06 and 07). Therefore, detection of the viral RNA in the BALF might be necessary for the monitoring of viral shedding, especially the patients in severe conditions.

Studies have shown that 2019-nCoV could utilize Angiotensin-converting enzyme 2 (ACE2) as the receptor to infect the host as SARS-CoV did ^11,13^. Interestingly, BALF samples from the severe cases possessed 100% positive rate, while in contrast, no viral RNAs were detected in the three BALF samples from mild cases. Although the sample size was small, it also suggests that the viral distribution is associated with diseases severity. More importantly, why the viruses in some individuals retained in the upper respiratory tract merits further elucidation.

Our study also has some limitations. Firstly, all the included cases were CDC-confirmed NCP patients, which may result in bias of sample selection. Meanwhile, studies have shown that 2019-nCoV caused asymptomatic infection in some individuals ^2,7,8^, and information of such patients is missing in our study. Secondly, most of samples were collected after antiviral treatment, which may influence the viral shedding. Third, the number of BALF samples was limited, especially for the mild cases. So, it is necessary to include more BALF samples to draw a more precise conclusion on the differences of viral shedding between the severe and mild cases.

In conclusion, sputum is most accurate for laboratory diagnosis of NCP, followed by nasal swabs, while throat swabs was not recommended for the diagnosis. Detection of viral RNAs in BLAF is necessary for the diagnosis and monitoring of viruses in severe cases. In addition, CT scan could serve as an important make up for the diagnosis of NCP. The NCP cases are rapidly increasing, and we hope that this study could provide useful information for the diagnosis and control of the 2019-nCoV infection.

## Data Availability

All data generated or used during the study have been presented in the submitted article.

## CONTRIBUTOR

YL, LL, ZZ, YY contributed to the study design. FW, JY, JL, MZ, ZW, LP, WW, JL contributed to the collection of clinical specimens. LX, JW, HZ, KF, QY, ML, JZ contributed to experiments and data collection. YY, MY and CS contributed to the data analysis. YY, MY, CS and WG contributed to the manuscript preparation.

## ACKNOWLEDGMENTS

This work was supported by the National Science and Technology Major Project (2017ZX10103011, 2017ZX10204401, 2018ZX10711001), Sanming Project of Medicine in Shenzhen (SZSM201412003, SZSM201512005) and China Postdoctoral Science Foundation (2019T120147, 2019M660836).

